# Algorithm for a multidisciplinary diagnostic and therapeutic approach to abdominal lymphatic malformations

**DOI:** 10.1101/2025.02.15.25322012

**Authors:** Annegret Holm, Dorian Marckmann, Jerry Wei Heng Tan, Susanne Lagrèze, Wibke Uller, Alexander Puzik, Rainer Misgeld, Claudia Blattmann, Stefan Fichtner-Feigl, Charlotte M. Niemeyer, Friedrich G. Kapp

## Abstract

**Objective:** Abdominal lymphatic malformations (ALM) are benign macro-, microcystic or mixed lesions that originate from defects in vascular development. Depending on size and location, clinical symptoms vary from asymptomatic to potentially life-threatening. Standardized diagnostic and therapeutic guidelines for ALM do not exist. Current literature focuses on imaging characteristics rather than symptoms for indication of invasive treatment strategies, including sclerotherapy and surgery. Sirolimus is evolving as a medical treatment option, while a watchful waiting approach for asymptomatic and low-risk ALM patients is not yet established as a conservative strategy.

**Methods:** We retrospectively characterized a multicenter cohort of 21 ALM treated 1996 - 2024. Items included clinical symptoms, imaging findings, and management strategies, including resection, sclerotherapy, sirolimus or a watchful waiting approach.

**Results:** ALM patients were categorized based on symptoms and lesion characteristics with potentially associated complications depending on lesion size and anatomical location: 9/21 (43%) patients were observed in the asymptomatic and low-risk group with regular clinical and radiological follow-up intervals. Acutely symptomatic (7/21; 33%), symptomatic or high-risk patients (5/21; 24%) underwent resection (10/21; 48%), sclerotherapy (1/21, 5%), or were treated with sirolimus (1/21, 5%). All patients had favorable long-term outcomes.

**Conclusions:** We developed a multidisciplinary diagnostic and therapeutic algorithm for ALM based on clinical presentation and lesion characteristics to ensure a standardized, yet patient-centered symptom- and risk-based approach for the most beneficial treatment and outcome. Importantly, our study is the first to demonstrate that watchful waiting is an adequate strategy for asymptomatic ALM patients with a low anticipated risk for complications. Validation of our proposed algorithm is warranted in a prospective study.

## 1. INTRODUCTION

The lymphatic system is essential for immunosurveillance and tissue fluid homeostasis. It comprises a dense vascular network of blind-ended lymphatic capillaries that drain interstitial fluid into valved collecting ducts for eventual return into the venous system.^1,2^ Anomalies within the lymphatic system arise due to benign defects during vascular development. Advances in understanding the genetic origin of these defects have helped to identify somatic activating mutations in PI3K/AKT/mTORC1 and RAS/MEK/ERK signaling pathway in the pathogenesis of lymphatic anomalies.^3–5^ Lymphatic anomalies are classified into simple and complex lesions.^6^ Simple, most often solitary lymphatic malformations (LM) are further subdivided based on their morphology into macrocystic, microcystic or mixed lesion subtypes and classically harbor somatic activating mutations in *PIK3CA*.^4,7^ In contrast, complex lymphatic anomalies (CLA) manifest in a distinct multifocal fashion and may include several tissues and central collecting ducts. These conditions comprise a continuum of complex lymphatic phenotypes including generalized lymphatic anomaly (GLA), central conducting lymphatic anomaly (CCLA), Gorham-Stout disease (GSD), and kaposiform lymphangiomatosis (KLA) with causative genetic variants in both PI3K and RAS signaling. Simple LM can form at any site; their occurrence in the abdominal lymphatic system is of particular interest since half of the lymph fluid is formed in the viscera and the system is essential for resorption of triglycerides transported with chylomicrons.^8,9^. Abdominal lymphatic malformations (ALM) account for approximately 5% of all lymphatic anomalies. Clinical presentation encompasses incidentally found, asymptomatic and symptomatic lesions that may result in life-threatening conditions in severe cases involving obstruction of adjacent anatomical structures with potential ischemia, hemodynamically relevant lesion bleeding as well as infection. ^10,11^. Besides the wide range of clinical presentations of ALM patients, the diagnosis of ALM is further challenged by the variety of possible benign and malignant differential diagnoses that may necessitate immediate action. Several case reports have reported on life-threatening courses of ALM. Diagnosis and treatment indication and modality is most often based on imaging findings and LM subtype characteristics: sclerotherapy is most often the treatment of choice for macrocystic and surgery for microcystic lesions in these case reports.^12–16^ Medical treatment with rapamycin (sirolimus), a mammalian target of rapamycin (mTOR) inhibitor, is emerging, preferentially for microcystic lesions.^16–18^ Some of the aforementioned studies consider symptom-based therapeutic strategies including a watch and wait approach. ^18,19^. However, no standardized approach to the diagnosis and treatment of ALM patients currently exists that takes into consideration the wide spectrum of clinical presentations, possible differential diagnoses, imaging findings with lesion characteristics, different treatment modalities, and associated risks. In this retrospective case series, we sought to develop a patient-centered, risk-stratified diagnostic and therapeutic algorithm with a focus on clinical presentation as well as lesion- and treatment-associated risks for optimal long-term outcomes for ALM patients.

## 2. MATERIALS AND METHODS

### Patients and data collection

Data of 21 ALM patients were collected retrospectively from four German centers for Pediatric Hematology/Oncology from 1996 to 2024 with approval by the Ethical Committee of the University Medical Center Freiburg in place (project number 464/19). Diagnosis of ALM was made either by indicative radiologic findings on MRI and in part by histopathologic analysis. All patients were contacted for a follow-up interview regarding their current condition with potential abdominal complaints. ALM lesions were classified as macrocystic, microcystic or mixed.^6^ Anatomical localization was categorized as originating from the mesenterium, the omentum, the retroperitoneal space or as unspecified in case the exact anatomical attribution was not possible. Patients were further categorized based on clinical presentation: Patients with acute symptoms (e.g., ileus and bowel ischemia) were classified as *symptomatic acute*. Symptomatic patients with recurrent episodes of pain, gradual obstruction of adjacent structures etc. as well as asymptomatic lesions associated with risk factors prone for potential complications were categorized as *symptomatic* or *high risk*. The third group consisted of patients with incidental findings of asymptomatic ALM lesions with low anticipated risk for complications were classified as *asymptomatic* and *low risk*. Treatment depending on category included resection, sclerotherapy or medical therapy with sirolimus or an observational approach with imaging controls. Patient follow-up was conducted as clinically indicated. Patients with typical differential diagnoses for ALM were added in Table 5 and Figure 3.

### Outcomes

Patient outcomes were defined as: complete remission if follow-up imaging showed no residues of the ALM lesion and no relevant clinical symptoms; partial response if imaging showed a decrease in size with no or decrease in clinical symptoms; stable disease in asymptomatic patients in the watchful waiting category with no significant increase in size in the imaging controls.

### Statistical Analysis/Descriptive analysis/Figures

Data were analyzed and plotted using GraphPad Prism 10.1.0 and Microsoft Excel. The schematic (Figure 4) was created with *BioRender*.

## 3. RESULTS

*Patient characteristics:* Twenty-one patients presenting with an ALM between 1996 and 2024 were included in this study. The gender distribution was according to previous reports with a larger proportion of male patients (14/21; 67%). The median age at diagnosis was 5 years (range from prenatally to 49 years), including adult patients with incidental findings in the asymptomatic group. Twelve patients (12/21; 58%) were in the range of 0 to 6 years; two patients (2/21; 10%) were diagnosed prenatally due to incidental finding in the routine prenatal ultrasounds. One patient received an additional fetal MRI for early evaluation, the other underwent further imaging after birth. Four patients (4/21; 19%) were older than 18 years (range 20 to 49 years) when diagnosed with an ALM. The median follow-up time was 3 years (range 1 - 26 years), the median age at last follow-up was 14 years (**Table 1**).

**Table 1.**
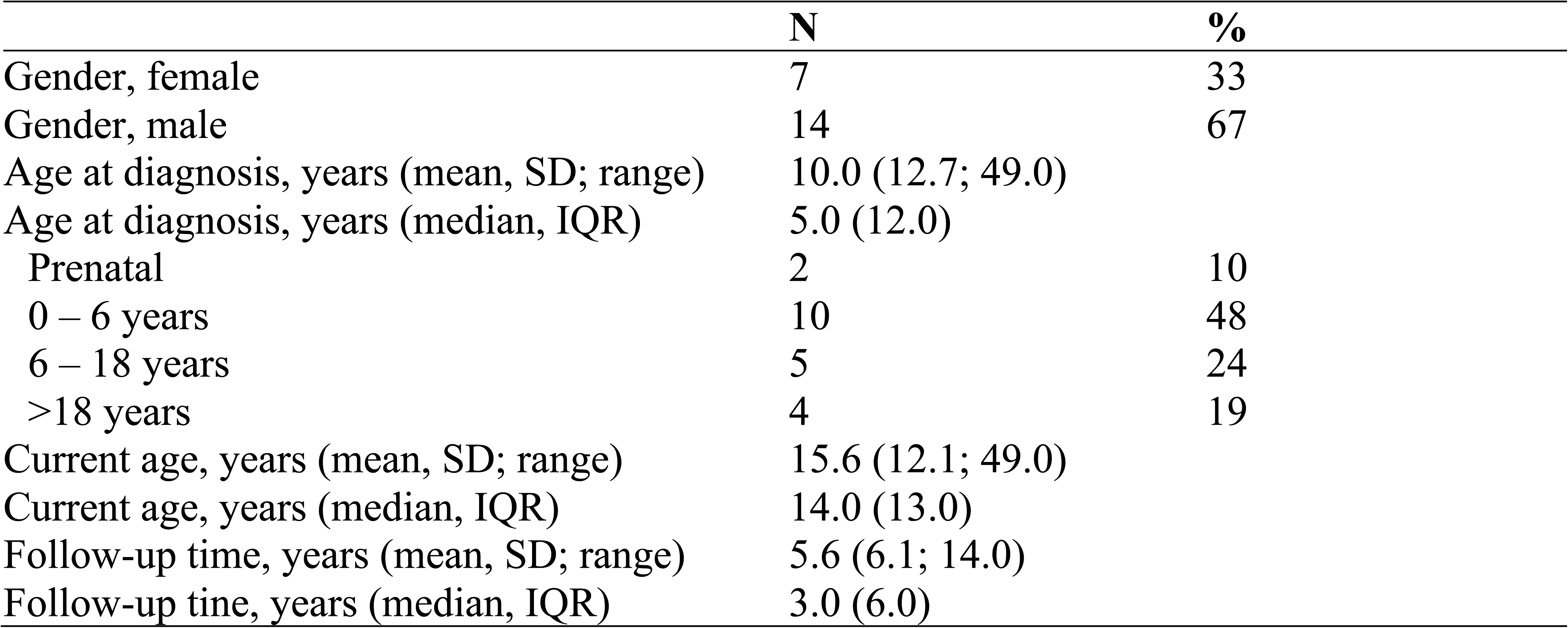
Patient Demographics (N = 21).

### Subtype and location of ALM

ALM were classified into the respective subtypes (macro- /microcystic/mixed) and attributed to an anatomical localization in the abdomen as described above. The majority of ALM lesions was classified as macrocystic (14/21; 67%) one as microcystic (1/21; 5%), and the remaining lesions (6/21; 29%) showed mixed macro- and microcystic components. Most of the ALMs originated from the mesenterium (9/21; 43%) or the omentum (2/21; 10%). The remaining lesions were either located in the retroperitoneal space (5/21; 24%) or classified as unspecified (5/21; 24%) due to the lack of a distinct separation from the surrounding tissue in either MRI or surgery (**Table 2**). Genetic testing was performed in two patients following surgery (2/21; 10%), revealing a hot spot mutation in exon 10 of *PIK3CA*, respectively.

**Table 2.** ALM Characteristics (N = 21).

|  | N | % |
| --- | --- | --- |
| LM Subtype |  |  |
| Macrocystic | 14 | 67 |
| Microcystic | 1 | 5 |
| Mixed | 6 | 29 |
| LM Location |  |  |
| Mesenteric | 9 | 43 |
| Retroperitoneal | 5 | 24 |
| Omental | 2 | 10 |
| Unspecified | 5 | 24 |
| Clinical Presentation at Diagnosis |  |  |
| Symptomatic | 11 | 52 |
| Asymptomatic | 10 | 48 |
| Classification |  |  |
| Symptomatic acute | 7 | 33 |
| Symptomatic or high risk | 5 | 24 |
| Asymptomatic and low risk | 9 | 43 |

### Clinical presentation of ALM

The patients presented initially either as symptomatic (11/21; 52%) or as asymptomatic with an incidental finding in imaging for a different indication (10/21; 48%). Seven *symptomatic acute* patients (7/21; 33%) presented with an acute abdomen requiring immediate exploratory laparotomy. Five patients (5/21; 24%) were classified as *symptomatic* or *high risk* based on size, location and growth dynamic of the lesion. The remaining nine patients (9/21; 43%) were classified as *asymptomatic* and *low risk* (**Table 2**). One initially asymptomatic patient developed symptoms during observation and hence managed and classified according to the *symptomatic* group with diagnostic workup and subsequent surgery (**patient 9, Table 4**). **Figure 1** shows MRI image examples and intraoperative findings of ALM patients of the different risk categories and respective treatment approaches.

**Figure 1.**
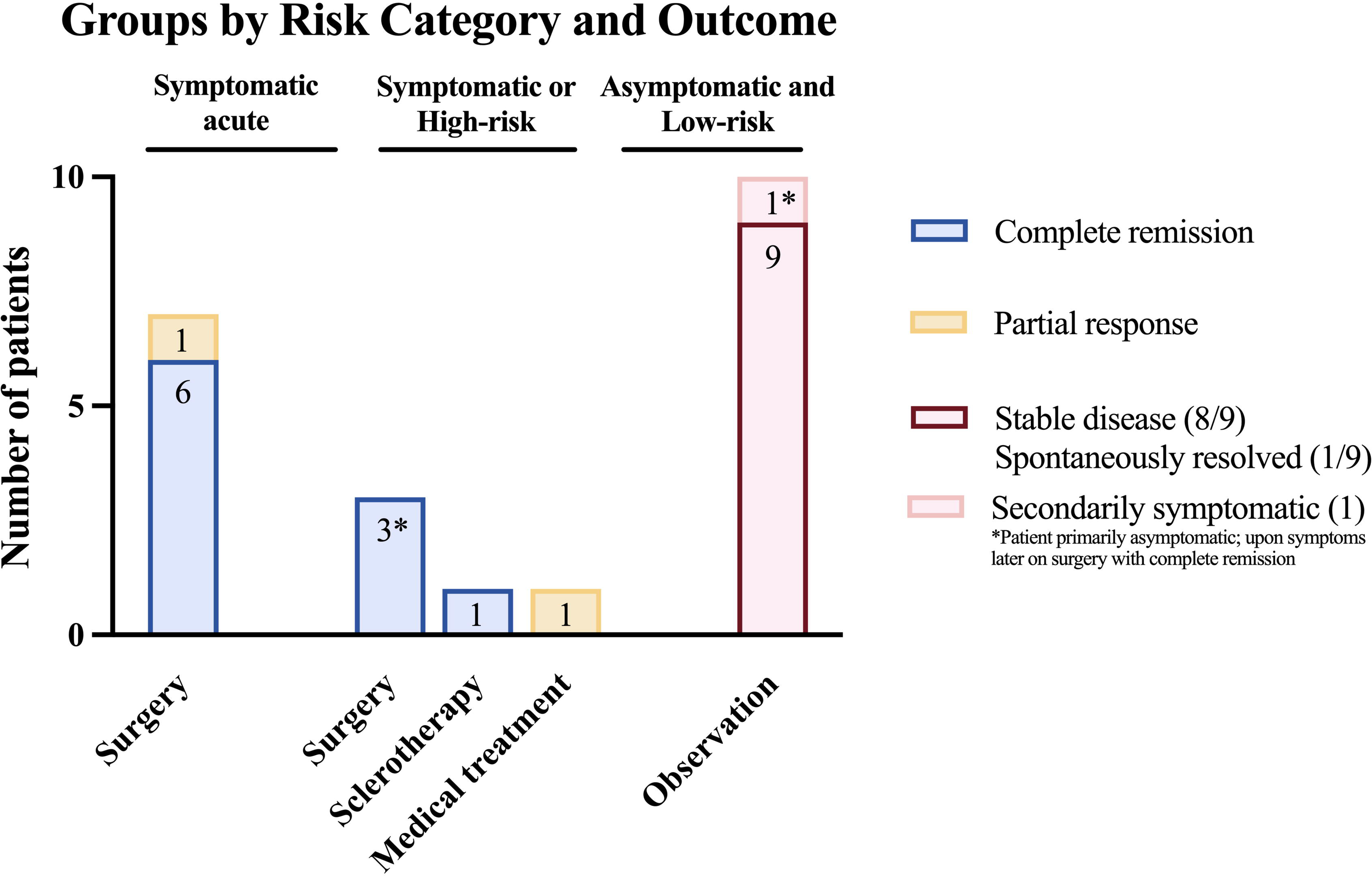
MRI and intraoperative findings for exemplary patients treated with different modalities. Attributed location and treatment modality as described in the panel.

### Outcomes

Patients were treated according to the recommendation of our multidisciplinary board for vascular anomalies consisting of Pediatric Hematologists/Oncologists, Diagnostic and Interventional Radiologists, Neuroradiologists, Pediatric, Maxillo-Facial, and Plastic Surgeons, among others. Treatment indication and modality was primarily based on clinical presentation and anticipated risk for complications. All patients presenting as *symptomatic acute* underwent emergency surgery (7/21; 33%) with complete resection of the ALM if possible. Most resected ALM were located in the mesenterium (5/7; 71%). One patient (patient 2, Table 4) underwent explorative laparotomy postnatally without resection of the ALM lesion; the patient underwent complete resection at age 16 years due to an ileus and experienced severe postoperative complications with anastomosis insufficiency, intestinal perforation necessitating several surgical revisions. One patient (patient 20, Table 4) - classified as partial response - with a large mesenteric ALM underwent incomplete resection and remains asymptomatic. Three of the five *symptomatic* or *high-risk* patients (5/21; 24%) underwent elective surgery. Overall, ten patients (10/21; 48%) underwent surgical resection with a complete remission in 9/10. All surgically treated patients had a favorable long-term outcome without relapse. 8/10 (80%) had a mesenteric ALM, one patient had an omental lesion; and one could not be unequivocally attributed anatomically (**Table 3**). One patient (1/21; 5%) in the *symptomatic* or *high-risk* group underwent sclerotherapy with doxycycline according to the recommendation of the multidisciplinary board and had a complete remission. On imaging the anatomic location of the lesion was anatomically not attributable (**Table 3**). One patient (1/21; 5%) in the *symptomatic* or *high-risk* category with a retroperitoneally located ALM was treated with sirolimus for 24 months. The patient is still on sirolimus and doing well with dose reduction as the lesion is significantly decreasing. An episode of neutropenia without infection resolved under dose reduction.

**Table 3.** Treatment Modalities.

|  | **N** | **%** |
| --- | --- | --- |
| Surgery | 10 | 48 |
| Mesenteric | 8/10 | 80 |
| Omental | 1/10 | 10 |
| Unspecified | 1/10 | 10 |
| Sclerotherapy | 1 | 5 |
| Unspecified | 1/1 | 100 |
| Medical Treatment | 1 | 5 |
| Retroperitoneal | 1/1 | 100 |
| Observation | 9 | 43 |
| Mesenteric | 1/9 | 11 |
| Omental | 1/9 | 11 |
| Retroperitoneal | 4/9 | 44 |
| Unspecified | 3/9 | 33 |

The patient group classified as *asymptomatic* and *low risk* (9/21; 43%) were closely observed with lesion-dependent intervals for clinical and imaging controls (ultrasound and/or MRI) according to the recommendation of our multidisciplinary board. One patient in the observational group with mixed mesenteric ALM developed intermittent pain and therefore underwent resection (therefore classified in the *symptomatic* group; this transition to another risk category is also highlighted in Figure 2); the patient has been asymptomatic ever since. All other patients remained asymptomatic and without a significant progress of the lesion on imaging. One patient experienced significant decrease in ALM lesion size and eventually complete regression in follow-up imaging controls (**Figure 1E, F and Patient 19, Table 4**). Anatomical location of the observational group varied, with one omental and mesenteric ALM each (1/9; 22%) and four retroperitoneally localized lesions (4/9; 44%). The remaining lesions (3/9; 33%) were anatomically not attributable in imaging (**Table 3**).

**Figure 2.**
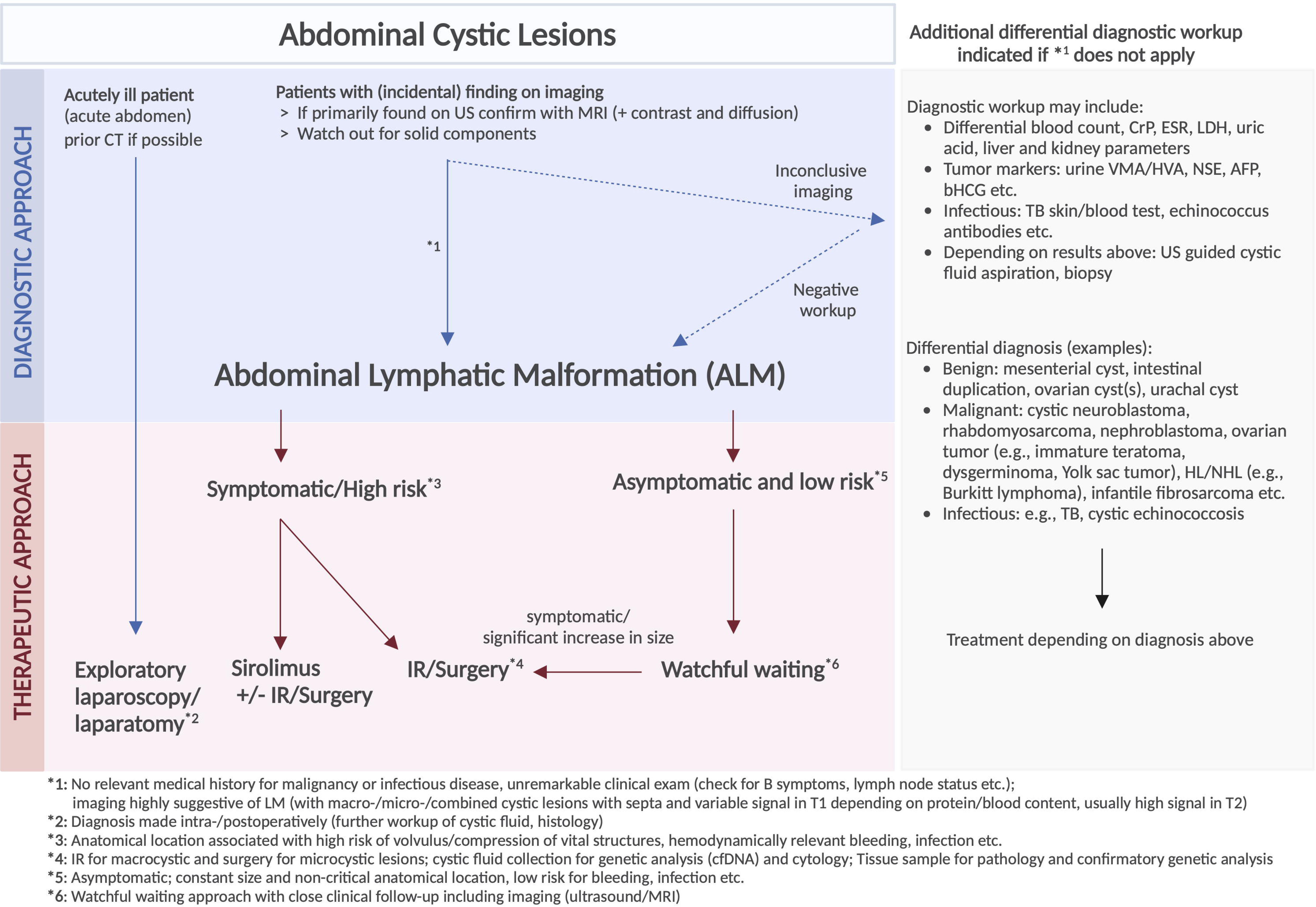
Symptom-based multimodal treatment and outcomes allocated to risk categories.

**Table 4.**
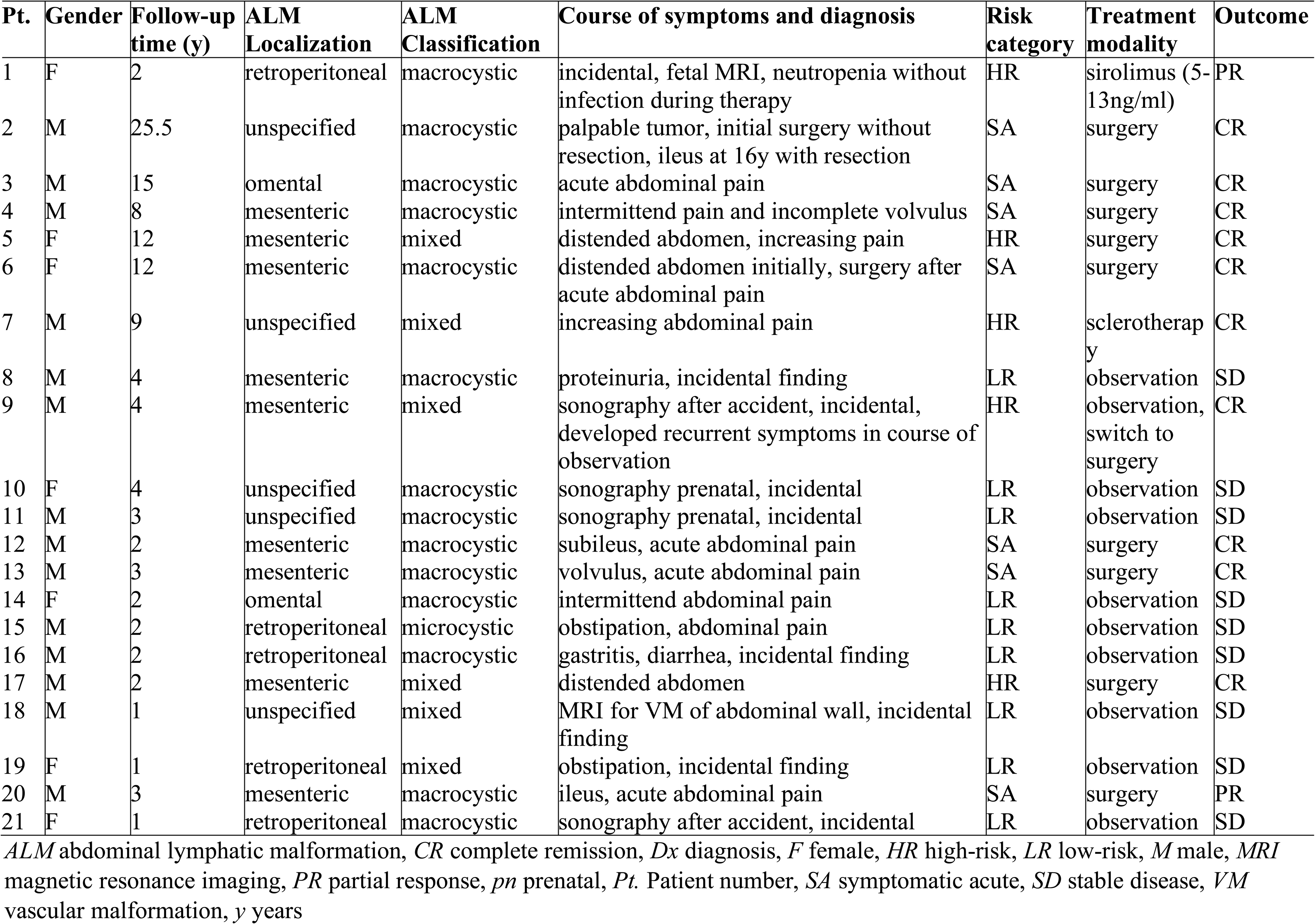
Summary of Patients (N = 21).

**Table 5.**
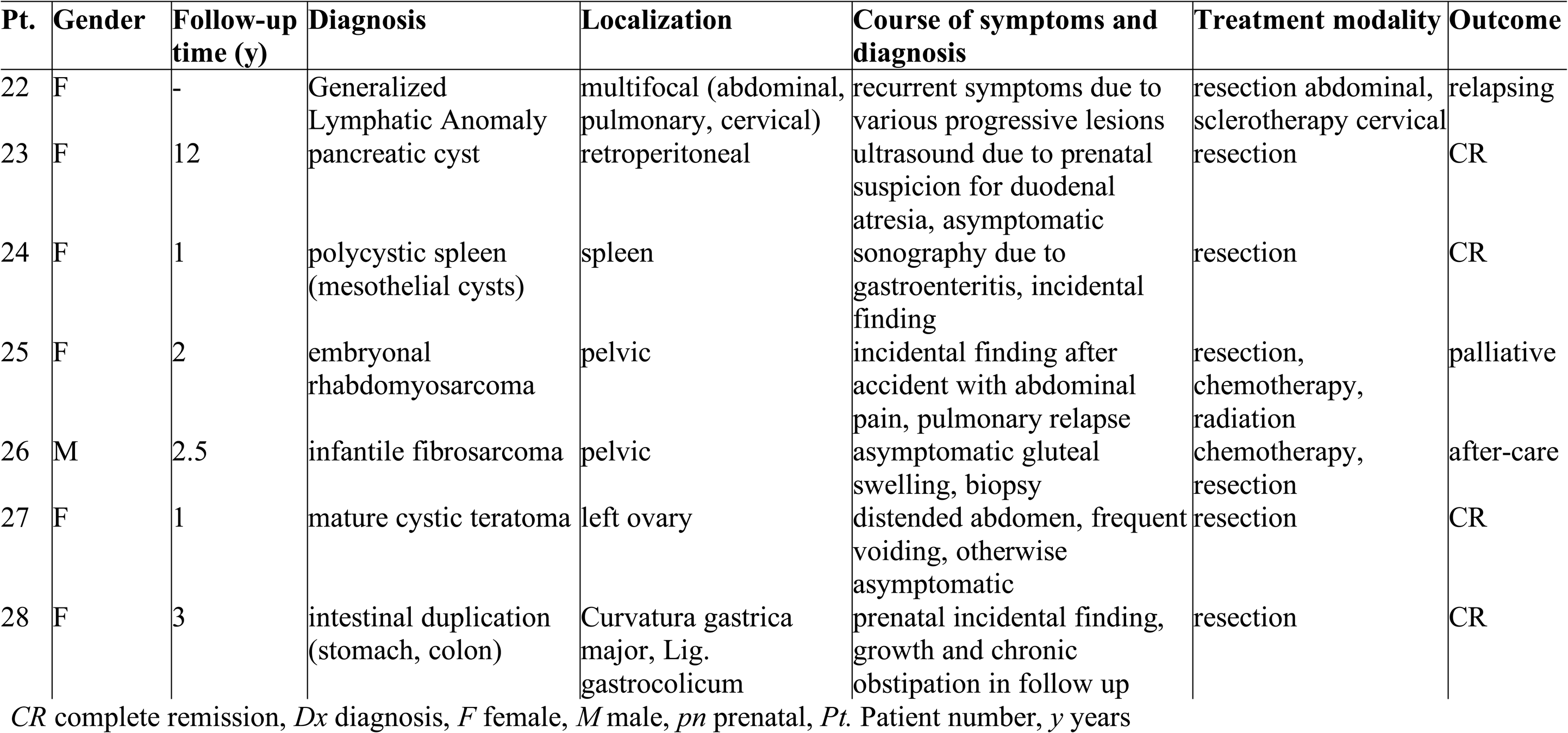
Summarized description of patients with possible differential diagnoses (N = 6).

In conclusion, all patients of our cohort had beneficial long-term outcomes. The risk-stratified treatment of choice in the *symptomatic* or *high-risk* cohort was surgery or sclerotherapy (for the latter using doxycycline) and sirolimus for lesions close to anatomically critical structures with high operative risk. Surgery and sclerotherapy were preferred for accessible macrocystic lesions, whereas sirolimus could be a good option for non-accessible lesions or lesions with microcystic parts. Observation was the approach of choice for *asymptomatic and low risk* patients and showed favorable outcomes. **Figure 2** exemplifies the varying treatment responses according to our risk classification.

## 4. DISCUSSION

In this study, we developed a complex diagnostic and therapeutic algorithm for the multidisciplinary management of patients with ALM. Guidelines for the diagnosis and treatment of ALM do not exist. Current strategies are based primarily on radiological lesion characteristics (micro-/macrocystic and mixed) rather than patient-specific symptoms and lesion-associated risks for complications depending on size and anatomical location.

Based on a retrospective, multicenter analysis of an ALM cohort of 21 patients, we aimed to offer a standardized approach to the diagnosis and treatment of ALM. Patient symptoms and potentially associated lesion complications - including compression of vital structures, volvulus, bleeding, and infection - were the main criteria for treatment indication and choice of treatment modality while taking relevant differential diagnoses into account.

To establish the diagnosis of an ALM, we propose that any stable patient with abdominal cystic lesions should obtain an MRI with contrast and diffusion-weighted sequences. Solid components on MRI and/or a clinical presentation suggestive of malignancy or an infectious origin (e.g., B symptoms, lymph node status) requires further differential diagnostic workup to ensure that other, most importantly malignant differential diagnoses are not missed; these may include Hodgkin or Non-Hodgkin lymphomas, cystic neuroblastoma, rhabdomyosarcoma, teratoma etc. Other benign or infectious differential diagnoses may comprise mesenterial cysts, intestinal duplications, ovarian cyst(s), mesenteric tuberculosis, cystic echinococcosis etc. (**Figures 3**, **4**).

**Figure 3.**
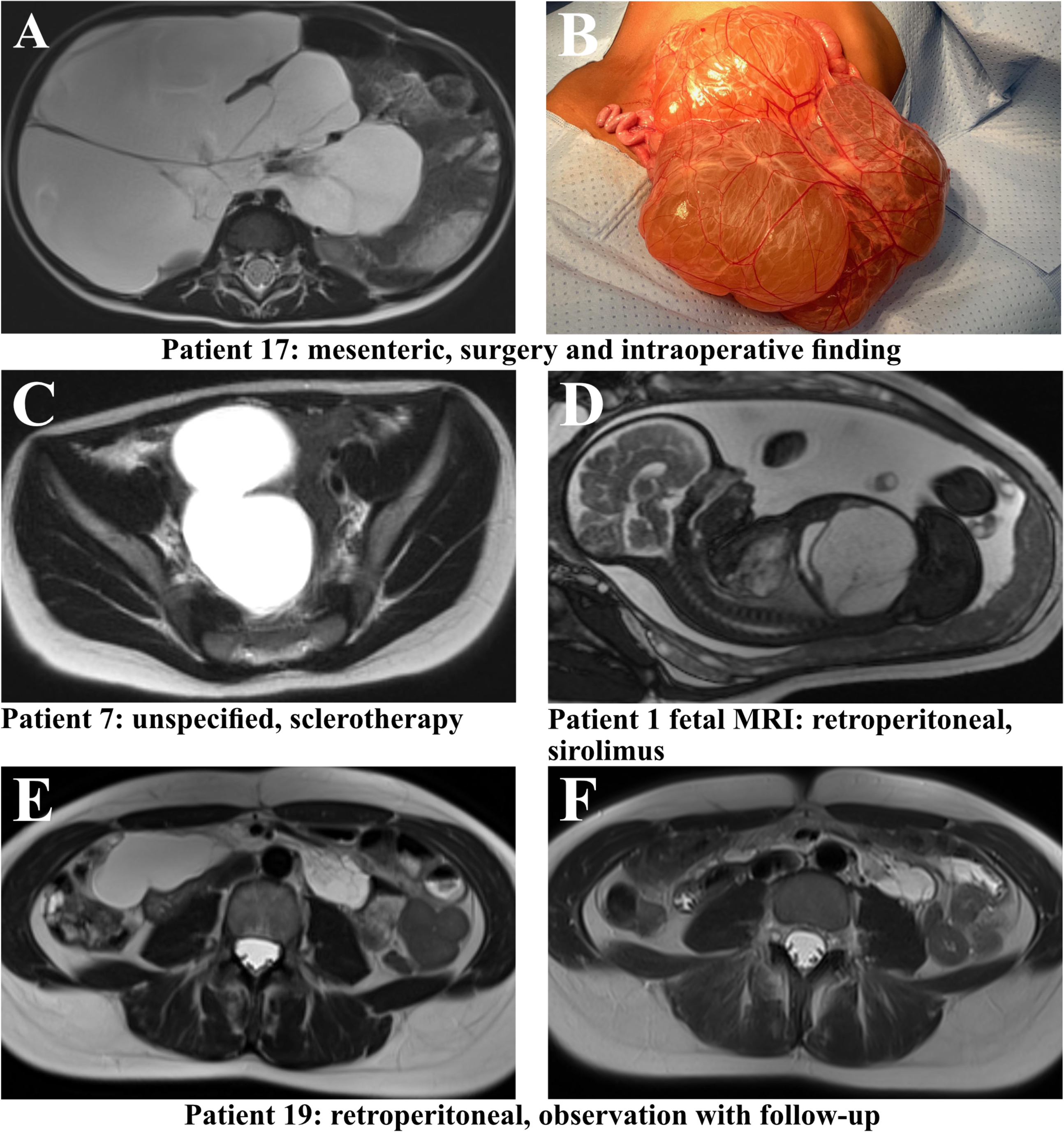
MRI of patients with potential differential diagnoses of ALM.

**Figure 4.**
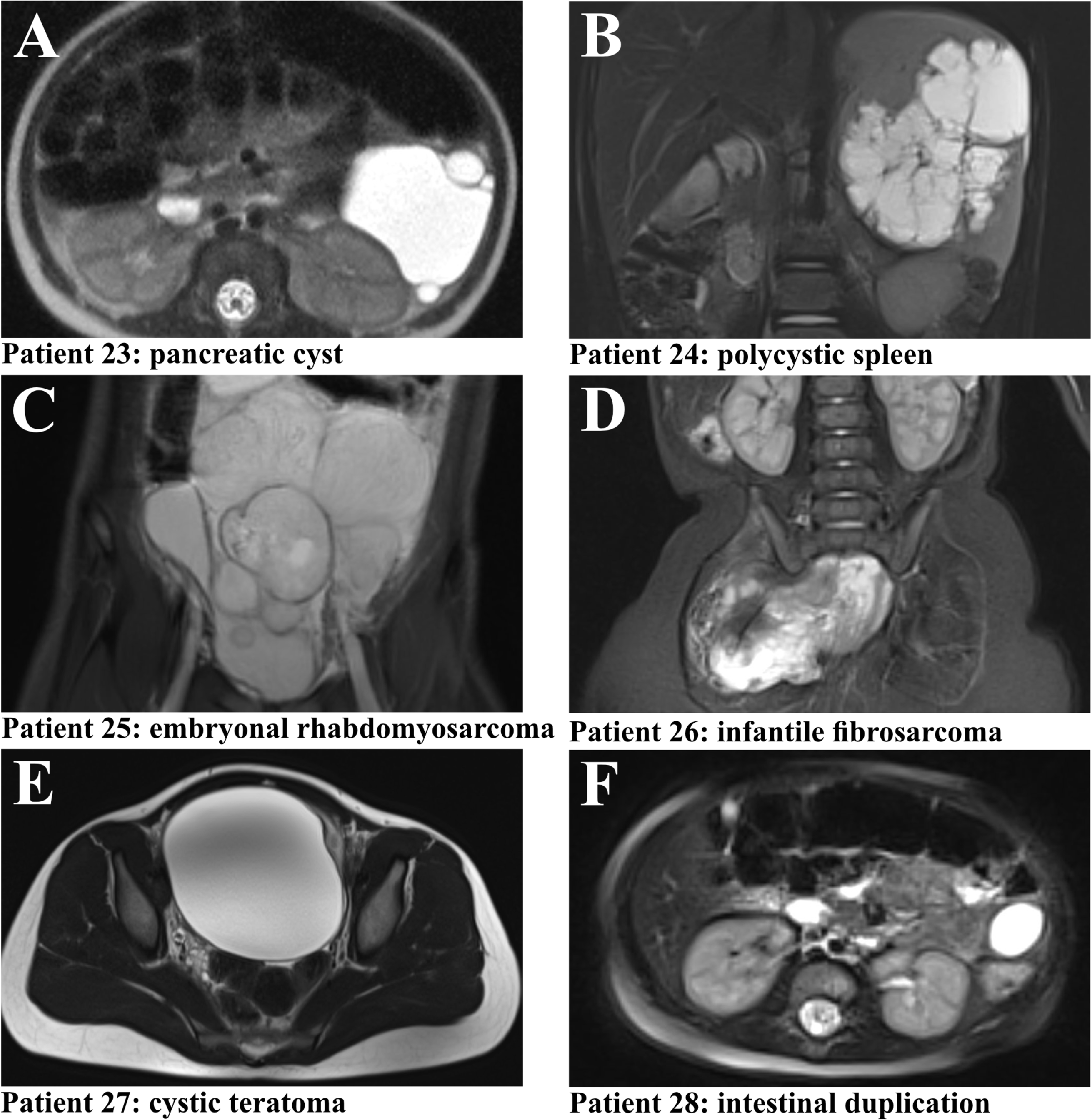
Algorithm for a multidisciplinary diagnostic and treatment approach for ALM.

Treatment strategies for ALM have been suggested by previous studies and include resection, sclerotherapy, and, more recently, mTOR inhibition with sirolimus. As is often the case in rare disease fields lacking standardized treatment algorithms, the respective center’s strongest expertise (surgery, interventional radiology, hematology/oncology) plays a role as to which treatment modality is prioritized. This treatment selection bias, however, may not result in the most beneficial treatment modality for each patient. Treatment algorithms have previously been suggested with a focus on sclerotherapy and/or surgery based on primary imaging findings, as mentioned in the introduction. Observation (watchful waiting) of primarily asymptomatic ALM patients with a low anticipated risk for complications is mentioned in the literature but not yet considered an established approach.^13,15,16,18–21^

Our study emphasizes the feasibility of a watchful waiting as a safe conservative approach in primarily asymptomatic and low-risk ALM patients. In fact, to the best of our knowledge, our study contributes the largest asymptomatic and low-risk cohort of ALM patients that did not undergo invasive procedures and demonstrated good long-term outcomes. Our study included 9/21(43%) long-term asymptomatic and low-risk patients. One patient with a mixed mesenteric LM was observed and experienced intermittent pain later on, which resolved following lesion resection. This case emphasizes the importance of close clinical and imaging follow-up intervals. The frequency depends on anticipated lesion complications based on size, anatomical location and growth dynamics. Reevaluation of the management approach is to be indicated in any patient that develops new symptoms and treatment modalities include resection, sclerotherapy, and medical therapy – a decision that is to be discussed in an interdisciplinary fashion. Intriguingly, the ALM in another patient showed nearly complete resolution over time **(Figure 1 E, F)** - an atypical feature in vascular malformations like LM. Benign vascular tumors such as infantile or congenital hemangiomas exhibit spontaneous involution, which remains mechanistically poorly understood. The continuous decrease in growth factors (VEGF-C) may play role, but further studies are warranted.^22,23^ The lesions of the remaining seven patients showed a constant size under a continued watchful waiting approach. In summary, these data suggest that watchful waiting is a safe conservative approach to asymptomatic and low-risk ALM patients with good long-term outcomes.

In our algorithm, we further differentiate patients in the symptomatic and/or high-risk group as acutely ill and those with intermittent or mild symptoms and/or a high-risk for associated lesion complications as outlined above. The acutely ill patient, often presenting with an acute abdomen, is prompted to undergo immediate explorative laparoscopy or laparotomy, ideally with a prior CT depending on the clinical situation. Symptomatic and/or high-risk patients may undergo sclerotherapy or resection based on radiological lesion characteristics as outlined in previous studies.^12,13,16,18^ Macrocystic lesions are amendable to sclerotherapy, while microcystic lesions may best be resected. Medical treatment with sirolimus may be beneficial in a neoadjuvant/adjuvant approach especially in large microcystic/mixed lesions prior to or following surgery, however, this has not been evaluated so far for abdominal lymphatic malformations and requires multidisciplinary discussion. Primary monotherapy with sirolimus may be indicated in lesions that are in critical anatomical locations and challenging to access via sclerotherapy or surgery.

We suggest collecting cystic fluid and tissue during surgery or sclerotherapy for cytology and genetic analysis of cell-free DNA (cystic fluid) and for pathology and genetic analysis of genomic DNA (tissue). This may be beneficial especially in complex lesions to establish a rationale for potential targeted medical treatments later on. In our cohort, we tested for and confirmed a hotspot somatic *PIK3CA* mutation (c1624G>A, p.Glu542Lys in exon 10, VAF 4% and 4.5%, respectively) in two patients. Both presented with large symptomatic and high-risk mesenterial macrocystic/mixed ALMs that each necessitated a median laparotomy. Genetic testing was not performed in the remaining patients. Genetic testing is the standard of care at our centers today; it was not considered a routine workup until recently due to the recently advancing genetic understanding and precision medicine strategies for vascular anomalies.^24^ As is the case for LMs, we anticipate that most ALMs may harbor a hotspot somatic *PIK3CA* mutation. Whether or not the presence of a *PIK3CA* mutation may predict the characteristics of an ALM lesion, associated complications and potential outcomes requires standard genetic testing in all ALM patients and thus further investigation.

Limitations of our study include the small patient cohort size, its retrospective nature, the relatively small number of patients in each treatment category (surgery, sclerotherapy and/or sirolimus), and the lack of genetic testing in most patients. These limitations are inherent to the rarity of ALM and the currently evolving treatment options. Nonetheless, our study provides valuable insights, highlights the safety of a watchful waiting approach instead of invasive or experimental treatments in low-risk patients, and may lay the foundation for future prospective studies, that may be conducted in recently established reference networks.

## 5. CONCLUSIONS

In conclusion, our study represents the first comprehensive algorithm for the diagnosis and treatment of ALM, integrating both individual patient symptoms, lesion-specific characteristics with potentially associated complications. We demonstrate the safety and efficacy of a conservative watchful waiting approach for asymptomatic and low-risk ALMs, offering a new perspective for a significant proportion of ALM patients. Our diagnostic workup with the most relevant benign/malignant and infectious differential diagnoses (**Figure 4**) provides further guidance to ensure medical conditions requiring specific treatment are not disregarded. By incorporating a patient-centered and multidisciplinary diagnostic and therapeutic approach, our algorithm represents a significant step towards an individualized, risk-stratified management of ALM patients in a standardized manner. Future prospective studies may build upon these findings to further refine and validate this approach, ultimately improving outcomes for patients with this rare and in part challenging condition.

## Data Availability

All data produced in the present work are contained in the manuscript.

## Conflict of Interest Disclosures (includes financial disclosures)

The authors have no conflicts of interest to disclose.

## Funding/Support

AH was supported by the Deutsche Forschungsgemeinschaft (DFG, German Research Foundation, grant 458322953). FK was supported by the charity cycling tour “Tour der Hoffnung” and received funding from the German Federal Ministry of Education and Research (BMBF): EJPRD Joint Transnational Call 2023, NARRATIVE; FKZ 01GM2405. The center at the University Hospital of Freiburg is a member of the Vascular Anomalies Working Group (VASCA) of the European Reference Network for Rare Multisystemic Vascular Diseases (VASCERN) - Project ID: 769036.

## Abbreviations

(ALM): abdominal lymphatic malformations
(LM): lymphatic malformations
(CLA): complex lymphatic anomalies
(GLA): generalized lymphatic anomaly
(CCLA): central conducting lymphatic anomaly
(GSD): Gorham-Stout disease
(KLA): kaposiform lymphangiomatosis
(mTOR): mammalian target of rapamycin

## ARTICLE SUMMARY

We propose a risk-stratified algorithm for abdominal lymphatic malformations (ALM) with invasive and medical strategies in high-risk and an observational approach for asymptomatic, low-risk patients.

## WHAT’S KNOWN ON THIS SUBJECT

ALM are benign macro- and/or microcystic lesions that may lead to life-threatening complications depending on size and location. Treatment guidelines for ALM patients do not exist and current strategies are based on lesion characteristics rather than clinical presentation.

## WHAT THIS STUDY ADDS

We developed a multidisciplinary diagnostic and therapeutic algorithm for ALM patients following a retrospective patient cohort analysis. Our proposed algorithm may contribute to safe and efficient patient-centered strategies while taking important differential diagnoses into account.

## Contributors Statement Page

Dr Annegret Holm contributed to study design, supervision, patient data acquisition and curation as well as patient follow-up, data analysis and interpretation, patient care, drafting and revising the manuscript.

Dr Dorian Marckmann contributed to patient data acquisition, analysis and interpretation, patient care, drafting and revising the manuscript.

Mr Jerry Wei Heng Tan, BS contributed to data analysis and interpretation.

Dr Susanne Lagrèze, Dr Alexander Puzik, Dr Rainer Misgeld, Dr Claudia Blattmann contributed to patient care.

Prof Wibke Uller contributed to radiological assessment and patient care.

Prof Charlotte Niemeyer and Prof Stefan Fichtner-Feigl contributed resources.

Dr Friedrich Kapp contributed to study design, supervision, patient care, drafting and revising the manuscript.

All authors approved the final manuscript as submitted and agree to be accountable for all aspects of the work.

## Notes

### Competing Interest Statement

The authors have declared no competing interest.

### Funding Statement

This study was funded by the Deutsche Forschungsgemeinschaft (DFG, German Research Foundation, grant 458322953), and by the charity cycling tour Tour der Hoffnung and received funding from the German Federal Ministry of Education and Research (BMBF): EJPRD Joint Transnational Call 2023, NARRATIVE; FKZ 01GM2405.

### Author Declarations

Ethics committee of the University of Freiburg, Faculty of Medicine gave ethical approval for this work.

